# Proteome-Wide Antigen Discovery Reveals Compartment-Specific Humoral Responses in Coccidioidomycosis

**DOI:** 10.64898/2026.08.24.26361245

**Authors:** Christine Boutros, Colette Caspar, Krista M. McCutcheon, Ravi Dandekar, Kelsey C. Zorn, Colin Zamecnik, Chloe Gerungan, Akshay Sharathchandra, Sukhman Sidhu, Christina M. Homer, Mark Voorhies, Marguerite Robison, Debanjana Chakravarty, Melody P. Lun, Alexis V. Stephens, Marie Nearing, Ramona Abbatista, Charles Y. Chiu, Joseph L. DeRisi, Manish J. Butte, David Meya, David Boulware, Jeffrey D. Whitman, George R. Thompson, Satya Dandekar, Anita Sil, Michael R. Wilson

## Abstract

Coccidioidomycosis is a fungal infection of rising public health concern, with coccidioidal meningitis (CM) representing its most devastating manifestation. Diagnosis of CM remains challenging due to the limited sensitivity and technical demands of conventional cerebrospinal fluid (CSF) serology testing. To comprehensively characterize the humoral immune response across the full spectrum of coccidioidal disease, we designed and deployed a proteome-wide *Coccidioides* phage immunoprecipitation sequencing (PhIP-Seq) library tiling both *C. immitis* and *C. posadasii* proteomes. We profiled antibody reactivity in sera from 323 participants spanning six clinically defined disease severity categories, as well as CSF and matched serum from participants with confirmed CM (n=108) and *Coccidioides*-negative other neurologic disease (OND) controls (n=163). Serum profiling revealed a potential narrowing of the antigenic repertoire as disease severity increased, with subclinical participants mounting the broadest response (55 peptides from 55 proteins) compared to 3-7 peptides in symptomatic categories. Two proteins, spherule outer wall glycoprotein (SOWgp) and a previously uncharacterized Proline-rich Immunodominant Antigen (PIA1), emerged as immunodominant across disease categories. Enriched antigens were disproportionately proline-rich and repetitive, a structural feature associated with immunodominance in other pathogens. CSF profiling revealed a compartment-specific antibody signature in CM, with 94% of CSF-enriched seroreactive peptides absent from matched sera. To orthogonally validate these findings, we developed a five-antigen Luminex assay using SOWgp- and PIA1-derived peptides, achieving 100% sensitivity and 100% specificity in both a discovery cohort (CM n=20, OND n=20) and an independent, blinded validation cohort (34 CM and 36 OND CSF samples. These findings expand the repertoire of *Coccidioides* serological responses associated with disease severity and demonstrate that proteome-wide antibody discovery can be translated into a targeted, high-performance diagnostic platform.

## INTRODUCTION

Coccidioidomycosis is a rising threat of critical public health importance. From April 2023 through March 2024, California reported 10,519 cases, representing a 39% increase over the same period in the previous year.^1,2^ *Coccidioides spp.* is a fungus that regularly causes disease in immunocompetent individuals.^3^ Disseminated coccidioidomycosis (DCM) occurs in 1% of infections, and a third of these cases are fatal.^4^ Despite the central nervous system (CNS) being a typically well-protected area against infection, up to half of DCM patients develop coccidioidal meningitis (CM).^5,6^ Survivors require lifelong antifungal therapy and often require surgical placement of a ventriculoperitoneal shunt to manage chronically elevated intracranial pressures.^7,8^

The diagnosis of CM is limited by the performance and prolonged turnaround times associated with labor-intensive reference laboratory methods for cerebrospinal fluid (CSF) testing.^9,10^ The most widely used diagnostic assays are immunodiffusion (ID) and complement fixation (CF). ID has specificity exceeding 95% in serum, but false positives in CSF can occur with high serum antibody titers that passively migrate from serum to CSF.^9,11^ CF has higher specificity, but this test requires significant technical expertise and has had a historical false-negative rate of 17% to 41%.^9,11^ Enzyme-linked immunosorbent assays (ELISA) are commercially available for *Coccidioides* serology but do not demonstrate superior performance to CF or ID and are prone to false-positive results.^12,13^ The gold standard for definitive diagnosis remains direct detection of *Coccidioides spp.* from clinical specimens by culture or histopathology, although the sensitivity of these techniques is low and often require invasive procedures.^9,11,14,15^ Only one molecular PCR assay has been approved (GeneSTAT, St. George, UT, USA), and its use is limited to respiratory specimens. Its sensitivity is comparable to that of fungal culture,^16^ suggesting that sensitivity may be similar or lower in CSF because of the reduced fungal burden associated with CNS infection. These limitations can result in significant misses or delays in initiating therapy for a potentially fatal infection.

A complementary direct detection approach is the *Coccidioides* galactomannan antigen enzyme immunoassay (CAg EIA). Initial studies demonstrated antigenuria in the highest risk patients with most severe disease and a sensitivity of approximately 71% in urine and moderate sensitivity in serum, though cross-reactivity with other endemic mycoses has been noted.^17^ When applied to CSF in patients with suspected CM, the CAg EIA achieved a sensitivity of 93% and specificity of 100%, outperforming antibody-based CSF methods. Its use in combination with antibody testing has been recommended as the most sensitive approach to CM diagnosis.^18^ Despite this performance, the CAg EIA has not supplanted CF-based testing as the routine diagnostic benchmark, in part due to limited laboratory adoption and cross-reactivity concerns. Consequently, there is an urgent clinical need for rapid serologic biomarkers that offer increased throughput and improved sensitivity and specificity, allowing for earlier intervention and more effective real-time assessment of fungal burden across the disease spectrum.

Here, to enable a more agnostic and comprehensive assessment of disease stage- and body-compartment-specific immunodominant *Coccidioides spp.* antigens, we designed, built, and deployed a proteome-wide *Coccidioides spp.* phage display assay. Using this assay, we performed phage immunoprecipitation sequencing (PhIP-Seq)^19^ experiments to provide the first deep-resolution view of the humoral immune response across the full spectrum of coccidioidal disease, from subclinical exposure to disseminated disease, including CM.

## METHODS

### Cohorts

Participants were selected from clinical cohorts enrolled by University of California, San Francisco (UCSF), the University of California, Davis (UCD), the University of California, Los Angeles (UCLA), the Valley Fever Institute (VFI), and the Infectious Diseases Institute at Makerere University in Kampala, Uganda in collaboration with the University of Minnesota (UMN). Samples included CSF and serum collected from participants with confirmed coccidioidomycosis and from *Coccidioides*-negative controls. Non-endemic control sera were obtained from the New York Blood Bank (NYBB).

Coccidioidomycosis diagnosis was established using current clinical criteria, including CF titers, IgG/IgM serologic testing, and clinical evaluation. CM was defined by comprehensive testing including positive CSF and/or serum CF and/or immunodiffusion, culture and molecular detection (e.g. PCR or metagenomic next-generation sequencing) in conjunction with compatible clinical findings. CSF control samples were derived from participants who had chronic meningitis or other neurological diseases and tested negative for *Coccidioides* by ID and CF, or culture and molecular testing.

All samples were de-identified prior to analysis. Clinical metadata were collected when available and included age, sex, geographic location, infection status, and timing of CSF and/or serum sample collection relative to diagnosis. CF titers were available for a subset of participants. Exclusion criteria included insufficient sample volume (<200 µL) and prolonged storage (>5 years) prior to experimentation.

### Coccidioidomycosis subtype cohort (UCLA, NYBB) - Serum

To evaluate antigen reactivity across the clinical spectrum of coccidioidomycosis, we profiled sera from a cohort of participants with confirmed *Coccidioides spp.* infection across disease subtypes from UCLA (IRB-19-1109, IRB-21-1318). Participants were stratified into six clinically defined disease severity categories as previously described^21^: Category 0 (no prior coccidioidomycosis), Category 1 (unrecognized past infection), Category 2 (uncomplicated pulmonary disease), Category 3 (complicated pulmonary disease), Category 4 (extrapulmonary dissemination without CNS involvement), and Category 5 (extrapulmonary dissemination with CNS involvement). Categories were assigned by Infectious Diseases specialists using established clinical criteria incorporating symptom severity, radiologic findings, sites of dissemination, and treatment requirements. Non-endemic control sera from the NYBB were included as additional Category 0 samples.

### CM discovery cohort (UCD, UCSF) - CSF and matched serum

To identify CNS-specific antigens, we profiled CSF samples from participants with confirmed CM obtained through the UCD Center for Valley Fever Serology Laboratory (IRB# 1884442-8), along with coccidioidomycosis-negative CSF controls from the UCSF Neuroinflammatory Diseases (NID) study (IRB# 13-12236) and UMN (IRB# 1304M31361). When possible, matched sera were obtained.

### Luminex validation cohort - CSF

An independent validation cohort was assembled to evaluate candidate antigen performance using a custom Luminex assay. This cohort included CSF samples from participants with confirmed CM from UCSF and UCD, along with coccidioidomycosis-negative CSF controls from the UCSF NID study. Validation cohort samples were entirely non-overlapping with the CM CSF Discovery Cohort and with the discovery subset used for initial Luminex evaluation to avoid having a participant contribute samples to both cohorts.

### IRB approval

All studies were approved by the Institutional Review Boards at participating institutions, including UCSF, UCD, UMN, VFI, and UCLA. Written informed consent was obtained from participants where required.

### *Coccidioides sp.* phage display library construction and validation

To enable unbiased antigen discovery, we designed and constructed a proteome-wide phage display library representing *Coccidioides immitis* RS (GCF_000149335.2_ASM14933v2) and *Coccidioides posadasii* Silveira (GCA_018416015.2_ASM1841601v2)^22^ using protein sequences obtained from GenBank in March 2022. Additionally, putative novel *Coccidioides* proteins were included, predicted from transcripts not annotated in current *Coccidioides* genomes but detected in RNA-seq datasets spanning the entire lifecycle of *Coccidioides.*^23^ These novel transcripts were annotated using the first stage of the transcriptome assembler from Gilmore et al.,^24^ with modifications for single-end Illumina reads. Specifically, reads from Mandel et al^25^ and Figure 1 of Homer et al ^23^ were aligned to the *Coccidioides posadasii* Silveira reference using BOWTIE version 2.4.2^26^ with default parameters. Exons were defined as contiguous genomic regions with read coverage ≥ 50 reads/base over all samples. Transcripts were assembled from exons by joining all exons bridged by at least two reads, annotating the bridged regions as introns. Transcripts were considered novel if they had no same-strand overlap with genes annotated in the reference. For each novel transcript, a protein sequence was inferred as the largest open reading frame in the spliced transcript sequence, and protein sequences of fewer than 60 amino acids were discarded.

**Figure 1:**
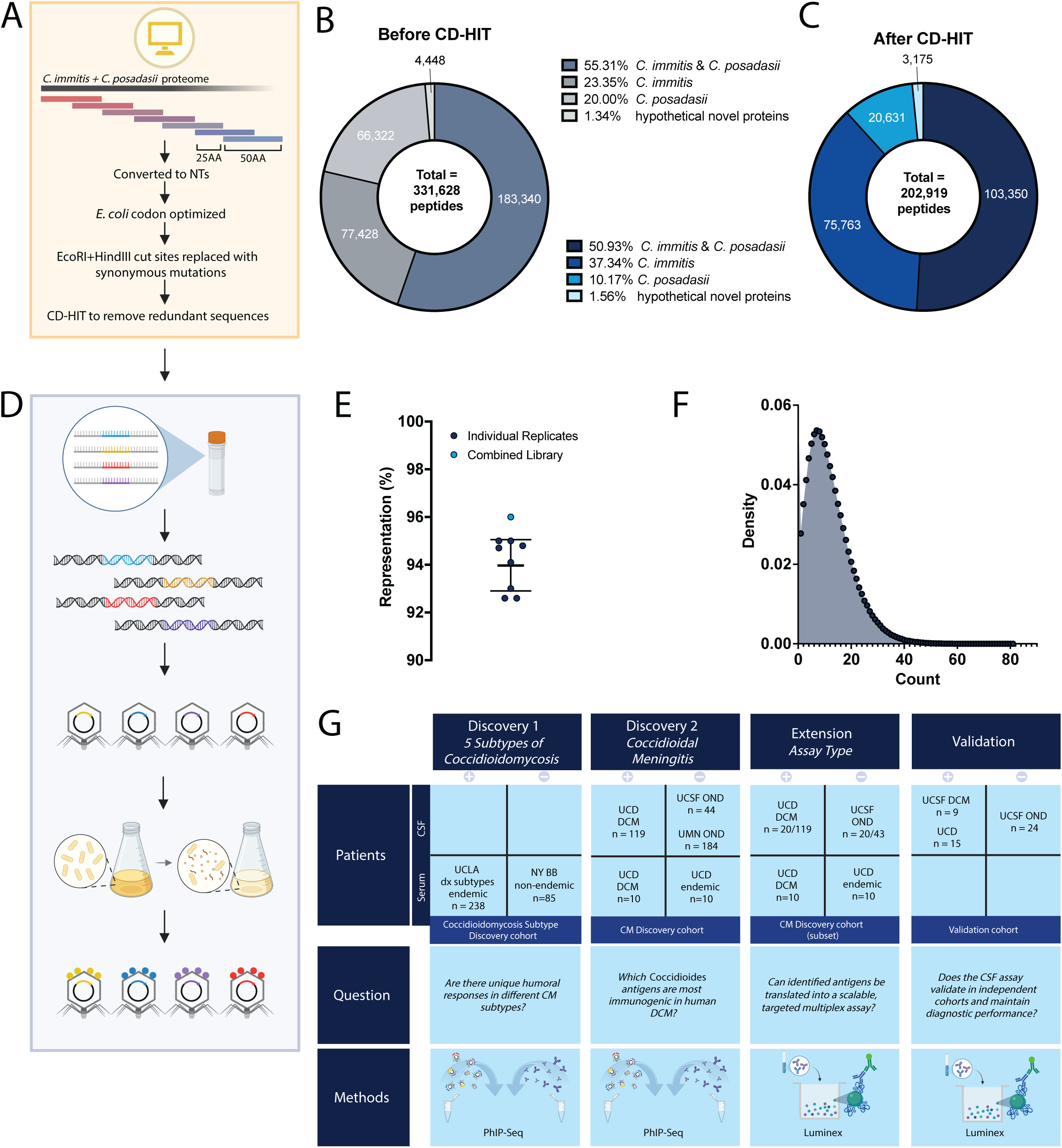
Design and validation of a proteome-wide *Coccidioides* PhIP-Seq library. **(A)** Schematic of library design. Protein sequences from *C. immitis* RS and *C. posadasii Silveira* were tiled into 50-amino acid peptides with 25-amino acid overlap to ensure full proteome coverage. **(B)** Donut chart displaying initial peptide library complexity (329,689 candidate peptides) by source genome prior to redundancy reduction. **(C)** Sequence clustering using CD-HIT2 at 95% amino acid identity reduced library size to 202,919 oligonucleotides. **(D)** Cloning of oligonucleotides into a T7 bacteriophage display system followed by amplification in *E. coli*. **(E)** Deep-sequencing-based quality control demonstrated that ∼96% of the designed peptides were represented in the final library. Final library titer reached 9 × 10¹ PFU/mL, corresponding to >100-fold coverage of the designed oligonucleotide pool. **(F)** Distribution of peptide abundance demonstrating near-normal representation and minimal cloning bias. **(G)** Experimental design schematic depicting four sequential stages: antigen discovery by PhIP-Seq across coccidioidomycosis subtypes and coccidioidal meningitis cohorts, targeted Luminex assay development in an extension cohort, and independent validation of CSF diagnostic performance.

Protein sequences were tiled into 50-amino acid (AA) peptides with 25-AA overlap to ensure full proteome coverage, generating 329,689 candidate peptide sequences (**Figure 1A, B**). All sequences were codon-optimized for *Escherichia coli* expression, and restriction enzyme recognition sites (HindIII and EcoRI) required for cloning were computationally identified and removed using synonymous codon substitutions.

To reduce redundancy, peptide sequences were clustered using CD-HIT2^27^ at 95% AA sequence similarity, resulting in a final library of 202,919 unique oligonucleotides (**Figure 1C**). These oligonucleotides were synthesized (Twist Bioscience, San Francisco, CA) and cloned into a T7 bacteriophage display system as previously described^28^ (**Figure 1D**). The amplified library was generated by infecting *E. coli* to enable large-scale phage production.

To assess library composition, the phage library was deep-sequenced before PhIP-Seq experiments. Paired-end reads were merged using Fast Length Adjustment of SHort reads (FLASH) software^29^ and aligned to the reference library using Bowtie2.^26^ Only exact sequence matches were included for downstream analysis.

### PhIP-Seq

#### Phage immunoprecipitation

Phage immunoprecipitation was performed through a previously published protocol.^20^ Briefly, immunoprecipitation was performed in duplicate using 2 μL of CSF or 1 μL of serum per participant, diluted 1:2 in 1x storage buffer and added to 150 μL of phage library (1:10 in storage media) per well, for a final sample dilution of 1:152, allowing antibodies to bind their corresponding peptide antigens displayed on phage. Antibody-phage immune complexes were captured using protein A/G magnetic beads. After washing to remove unbound phage, the bound phage were eluted and amplified through reinfection of *E. coli*. A second round of immunoprecipitation was performed to enrich antibody-bound phage before sequencing.

#### Sequencing library preparation and sequencing

Extracted phage DNA was PCR-amplified to generate indexed libraries for high-throughput sequencing. Each 25 µL reaction contained 12.5 µL 2X Phusion Hot Start HF Master Mix, 0.75 µL DMSO, 7 µL nuclease-free water, 1.25 µL pan-ome primer mix, 2.5 µL dual-index barcode primer, and 1 µL DNA template diluted 1:50. To prepare the template, 2 µL recovered lysate was first diluted in 8 µL molecular-grade water and then further diluted with 90 µL water.

PCR master mix was added to a fresh plate, followed by indexed barcode primers and diluted template. Reactions were mixed thoroughly, briefly centrifuged, and amplified using the following thermocycler conditions: initial denaturation at 95°C for 2 minutes; 5 cycles of 95°C for 30 seconds, 66°C for 30 seconds, and 72°C for 45 seconds; followed by 5 cycles of 95°C for 30 seconds, 64°C for 30 seconds, and 72°C for 45 seconds; followed by 20 cycles of 95°C for 30 seconds, 62°C for 30 seconds, and 72°C for 45 seconds; and a final extension at 72°C for 2 minutes, with a hold at 4°C. After amplification, 3 µL from each well was pooled by row to generate sequencing library pools. Pools were kept separate until post-PCR quality control (TapeStation and Qubit DNA concentration).

Amplification used a previously described “pan-ome” forward primer and indexed reverse primers containing 0- to 3-nucleotide stagger sequences to improve sequence complexity during Illumina sequencing.^30^ Dual index barcodes were incorporated during PCR to permit multiplexed sequencing. Indexed libraries were sequenced on an Illumina NovaSeq, and reads were used to identify antibody-enriched phage-displayed peptides.

The forward primer sequence was: ACACTCTTTCCCTACACGACGCTCTTCCGATCTAGTCAGGTGTGATGCTCGGGGATCC.

Reverse primer sequences were: GTGACTGGAGTTCAGACGTGTGCTCTTCCGATCTTAGTTACTCGAGTGCGGCCGC AAGC; GTGACTGGAGTTCAGACGTGTGCTCTTCCGATCTNTAGTTACTCGAGTGCGGCCG

CAAGC; GTGACTGGAGTTCAGACGTGTGCTCTTCCGATCTNNTAGTTACTCGAGTGCGGCC GCAAGC; and GTGACTGGAGTTCAGACGTGTGCTCTTCCGATCTNNNTAGTTACTCGAGTGCGGCCGCAAGC.

#### PhIP-Seq analysis

Fastq files were aligned to the reference *Coccidioides sp.* peptide library, and individual peptide counts were normalized to reads per 100,000 (RPK) by dividing by the sum of counts and multiplying by 100,000 to account for varying read depth. All statistical analyses were performed in Python (v3.11.7) using standard scientific computing libraries, including NumPy, pandas, SciPy, and scikit-learn.

### Identification of enriched seroreactive peptides

Peptide enrichment was quantified by comparing *Coccidioides*-positive biological samples to mock-IP controls and *Coccidioides*-negative biological samples. Samples were run in duplicate. For each peptide, fold-change enrichment was calculated relative to the mean signal observed in mock-IP controls. Z-scores were calculated by standardizing peptide-level signals using the global distribution of mock-IP peptides (mean and standard deviation across all mock-IP samples). Pairwise similarity of reactivity profiles across samples was assessed using Pearson correlation coefficients calculated on log-transformed RPK values. Correlation matrices were visualized as heatmaps.

For the Coccidioidomycosis Subtype Cohort, peptides were considered enriched if they met the following criteria: (i) ≥ 2-fold enrichment over mock-IP controls; (ii) Z-score ≥ 2; and (iii) total RPK ≥ 1.

For the CM Discovery Cohort, peptides were considered enriched if they met the following criteria: (i) ≥ 3-fold enrichment over mock-IP controls, (ii) Z-score ≥ 3, (iii) total RPK ≥ 10. Duplicates were then averaged.

A peptide was considered enriched for either the case or control group if it was enriched in at least 50% of samples. Protein-level enrichment was determined by summing the total number of enriched seroreactive peptide sequences mapping to the same protein.

### Structural and motif analysis of enriched seroreactive peptides

Repetitive sequence motifs within enriched *Coccidioides* proteins were identified using the Rapid Automatic Detection and Alignment of Repeats (RADAR) in protein sequences algorithm.^31^ For each protein, repeat elements identified by RADAR were summed to generate a cumulative repeat frequency. The cumulative repeat frequency per protein was compared between seroreactive and non-seroreactive proteins. To account for group size imbalance, non-seroreactive proteins were randomly sampled without replacement to match the number of seroreactive proteins, and a Mann-Whitney U test was applied to each resample. This procedure was repeated across 1,000 iterations.

### Luminex assay

CSF candidate antigens identified by PhIP-Seq were selected for orthogonal validation in a Luminex assay based on enrichment magnitude and reproducibility across samples. Selected antigens were coupled to Luminex beads according to the manufacturer’s protocol (Luminex Corp., Austin, TX). Five bovine serum albumin (BSA) consensus peptide conjugates were prepared, and experiments were run as previously described^32^. The net mean fluorescence intensity (MFI) for each peptide antigen was calculated for each sample by subtracting the MFI of that sample’s corresponding intra-assay BSA-background control and averaging across duplicate wells. Positivity thresholds for Luminex validation were empirically defined using MFI normalized to BSA-only bead background (MFI/background).

## RESULTS

### Study cohorts

#### Coccidioidomycosis Subtype Cohort (UCLA, NYBB)

A total of 238 serum samples were obtained from UCLA across the six disease severity groups as described in Methods: Category 0 (n=1), Category 1 (n=6), Category 2 (n=81), Category 3 (n=45), Category 4 (n=45), and Category 5 (n=60).^21^ The mean age was 47.0 years (SD 14.4; range 14-82), with a predominance of males (148/238, 62%). Eighty-five NYBB serum samples were grouped with Category 0 in combination with UCLA samples (n=86). Demographic data from the cohort are available in **Supplemental Table 1**. Demographic data were not available for the 85 non-endemic control sera from the NYBB. (**Supplemental Table 3**).

### CM discovery cohort (UCD, UCSF, UMN)

To identify CNS-specific antigens, we utilized the CM CSF Discovery Cohort, consisting of CSF samples from participants with confirmed CM (n=108 participants, contributing 117 samples) and *Coccidioides*-negative control CSF samples (n=43 participants).

Among the 108 CM-confirmed participants, the mean age was 46.1 years (SD 19.4; range 0-82), with a male predominance (79/108, 73%), consistent with the known epidemiology of DCM.^33^ Most CSF samples were obtained from California (102/117, 87%), with smaller contributions from Arizona (3/117, 3%), Indiana (1/117, 1%), and Washington (1/117, 1%). Geographic data were unavailable for a subset of samples (10/117, 8%) (**Supplemental Table 2**). In comparison, participants with *Coccidioides*-negative CSF had a mean age of 45.4 years (SD 24.5; range 1-81) and were 56% male (27/43). Diagnoses for the *Coccidioides*-negative control CSF samples included autoimmune conditions (n=21), viral meningitis or encephalitis (n=4; herpes simplex virus encephalitis, St. Louis encephalitis virus, West Nile virus, and suspected viral meningoencephalitis), bacterial meningitis or encephalitis (n=4; *Mycobacterium tuberculosis* n=2, Group A *Streptococcus* bacteremia with CNS involvement, and *Bordetella hinzii* brain abscess), fungal meningitis due to non-*Coccidioides* fungi (n=4; *Cryptococcus neoformans* n=2, *Aspergillus sp.* n=1, *Histoplasma capsulatum sp.* n=1), parasitic encephalitis (n=1; *Toxoplasma gondii*), neoplastic or paraneoplastic conditions (n=3), treatment-related neurotoxicity (n=3), and other neurological conditions of unclear etiology (n=3). An additional 120 samples were obtained from 120 participants with HIV-associated cryptococcal meningitis enrolled in Uganda, a *Coccidioides* non-endemic region. These participants were not explicitly tested for *Coccidioides* infection. Participants had a median age of 36 years (IQR 30-39) and were 58% male (35/60). All Ugandan participants were people living with HIV (60/60, 100%) with a median CD4 count of 31 cells/µL (IQR 13-66), and all had a diagnosis of cryptococcal meningitis (60/60, 100%).

For the CM participants, the mean time from CM diagnosis to CSF collection among samples with available metadata (n=108) was 271 days (SD 455; range 0 to 1679 days). Forty (37%) were collected on the day of diagnosis, 18 (17%) within 1-30 days after diagnosis, and 50 (46%) more than 30 days after diagnosis.

### Luminex validation cohort

Among the 18 UCSF CM participants (19 samples from 18 participants), of whom 10 had available metadata (note: one participant contributed two samples collected 95 days apart), the mean age was 34.7 years (SD 15.8; range 10-66), with a male predominance (8/10, 80%); all were from California. Among the 14 UCD CM participants (contributing 15 samples, including one participant with two longitudinal timepoints), the mean age was 39.2 years (SD 16.9; range 15-61) among the 13 with available age data, with a male predominance (10/14, 71%); 13 of 14 were from California, and 1 had an unknown geographic origin.

Among the 36 OND participants, 24 participants had available metadata. The mean age for these participants was 47.8 years (SD 17.6; range 10-81), with 11/24 (46%) male; the majority were from California (21/24, 88%), with single participants from Tennessee, Washington, and outside the United States. Diagnoses for the 24 OND participants included fungal meningitis due to non-*Coccidioides* pathogens (n=9; *Candida* spp. (n=3; *C. parapsilosis*, *C. dubliniensis*, and *Candida* sp.), *Aspergillus fumigatus* (n=1), *C. neoformans* (n=1), *H. capsulatum* (n=2), suspected endemic mycosis (n=1), and unidentified fungal pathogen in a patient with X-linked chronic granulomatous disorder (n=1)), neurosarcoidosis (n=5), autoimmune or inflammatory meningitis (n=2; including neuro-Behçet disease and idiopathic leptomeningitis/pachymeningitis considered likely autoimmune), idiopathic chronic meningitis of unclear etiology (n=6), suspected indolent bacterial meningitis (n=1), and nucleotide-binding domain, leucine-rich repeat, and pyrin domain-containing protein 3 (NLRP3)-associated autoinflammatory meningitis (n=1). The validation cohort additionally included 12 de-identified CSF samples with confirmed *Coccidioides* infection, but without additional clinical metadata.

### “Pan-*Coccidioides*” phage display library development

To confirm the “pan-*Coccidioides*” phage display library represented the designed library, we first determined the titer of the cloned and phage-packaged product. The library achieved a titer of 9 × 10¹ PFU/mL, corresponding to at least an estimated 100-fold coverage of the designed library. A post hoc review of the oligo library identified incomplete computational removal of HindIII restriction sites during the design stage. Despite this, deep sequencing of the library revealed that ∼96% of the designed peptides were present in the cloned library, with an average sequencing depth of approximately 1 million reads per sample. All samples were run in duplicate (**Figure 1E**). The frequency and distribution of peptide representation exhibited a near-normal distribution (**Figure 1F**), indicating minimal representation bias.

### *Coccidioides* PhIP-Seq reveals immunological signatures specific for disease severity

To comprehensively characterize the serological response across the coccidioidomycosis disease spectrum, we performed PhIP-Seq on serum samples (n=237 cases and n=86 controls) from the Coccidioidomycosis Subtype Cohort (**Figure 2A**). Pearson correlation analysis showed little within-group concordance (**Figure 2B**), indicating substantial inter-individual heterogeneity in the antibody response in serum. Despite screening the *Coccidioides* proteome, seroreactivity was remarkably sparse, with seroreactive peptides in serum mapping to only 55 proteins out of the approximately 7,000-10,000 proteins encoded by the fungal genome. Thus, the circulating serum antibody response to *Coccidioides* appears to target a relatively restricted set of antigens, with considerable variation in antigen recognition between individuals. Within this limited seroreactive landscape, enrichment analysis revealed distinct reactivity patterns in serum across disease categories **(Figure 2C)**, with enrichment mapping to multiple *Coccidioides* proteins, including the known extracellular antigen SOWgp^34–36^ (CIMG_04613/CPSG_06466) and an uncharacterized protein, here termed Proline-rich Immunodominant Antigen 1 (PIA1) (CIMG_05576/CPSG_05795). This protein was recently reported to be seroreactive in immunized or *Coccidioides*-infected dogs.^37^

**Figure 2:**
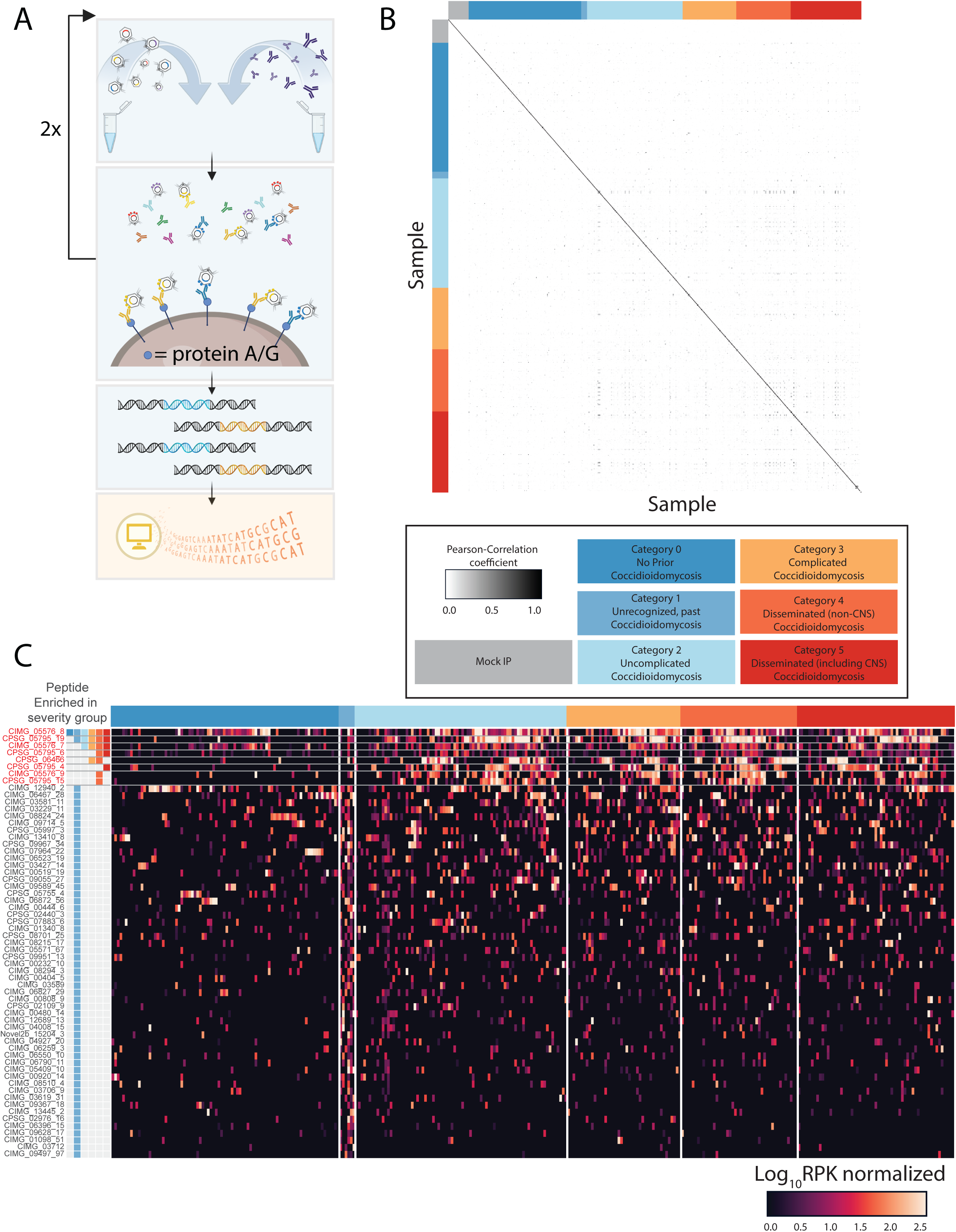
Serum antibody reactivity profiles across clinically defined coccidioidomycosis categories. **(A)** PhIP-Seq workflow: the T7 phage display library is incubated with human serum/CSF twice sequentially to enrich antibody-bound phage, which are then captured using protein A/G beads. Bound phage are eluted, and inserts are amplified and subjected to next-generation sequencing for downstream quantification of peptide enrichment. **(B)** Pairwise Pearson correlation matrix of log-transformed RPK values demonstrating low within-disease category concordance across serum samples, consistent with heterogeneous antibody responses. **(C)** Heatmap of enriched seroreactive peptides across disease categories following application of enrichment thresholds (≥2-fold over mock-IP, Z-score ≥2, RPK ≥1, present in ≥50% of samples per group).

Reactivity to PIA1 fragment 8 (AA200-250) was detected in all serum categories, including the coccidioidomycosis-negative cases (i.e., Disease Category 0), suggesting that it is not a specific marker of *Coccidioides* exposure. PIA1 fragment 19 (AA475-525) was enriched in categories 1-5 but absent in category 0, suggesting an antigen specific for *Coccidioides* exposure. PIA1 fragment 7 (AA175-225) was observed in categories 2-5 (i.e., symptomatic disease), whereas reactivity to SOWgp fragment 1 (AA1-50) was restricted to categories 3-4 (i.e., complicated or disseminated disease without CNS involvement).

The unrecognized previous infection group (Category 1) exhibited a diverse serological profile, with 55 total enriched seroreactive peptides in serum from 55 separate proteins, of which 53 (96.4%) were unique to this group. In contrast, symptomatic disease groups (categories 2-5) demonstrated convergent and narrower immune responses (3 peptides enriched in category 2, 4 peptides enriched in category 3, 7 peptides enriched in category 4, and 5 peptides enriched in category 5). Category 2 peptides were a subset of those in category 3, with category 3 additionally enriching SOWgp fragment 1 (AA1-50). PIA1 was the most common immunogenic protein. Extrapulmonary disseminated disease without CNS involvement (Category 4) was enriched for multiple PIA1 fragments, including two not observed in any other category: PIA1 fragment 9 (AA225-275) and PIA1 fragment 15 (AA375-425). CM participants (Category 5) demonstrated a unique serum signature, with peptide PIA1 fragment 4 (AA100-150) present exclusively in this category.

### *Coccidioides* PhIP-Seq identifies compartment-specific reactivities in the CM discovery cohort

We next explored the antibody profiles in the CSF of patients with CM to understand whether there was a humoral immune response unique to this anatomic compartment. Pearson correlation analysis demonstrated strong concordance among CM CSF samples, suggesting homogeneity in the CNS humoral response in CM CSF (**Figure 3A**). All CM-enriched seroreactive peptides had minimal reactivity in OND CSF controls (p-value <0.01; **Figure 3B**). The final dataset comprised 49 enriched seroreactive peptides derived from 9 distinct proteins across both *Coccidioides* species: PIA1, SOWgp, a Ph domain-like superfamily protein, a ubiquinol cytochrome c reductase complex protein, zinc finger protein 58, two orthologous guanyl nucleotide exchange factors, 3-oxoacyl-carrier protein reductase, and two non-orthologous hypothetical proteins. (**Supplemental Table 4**, **Figure 3C**).

**Figure 3:**
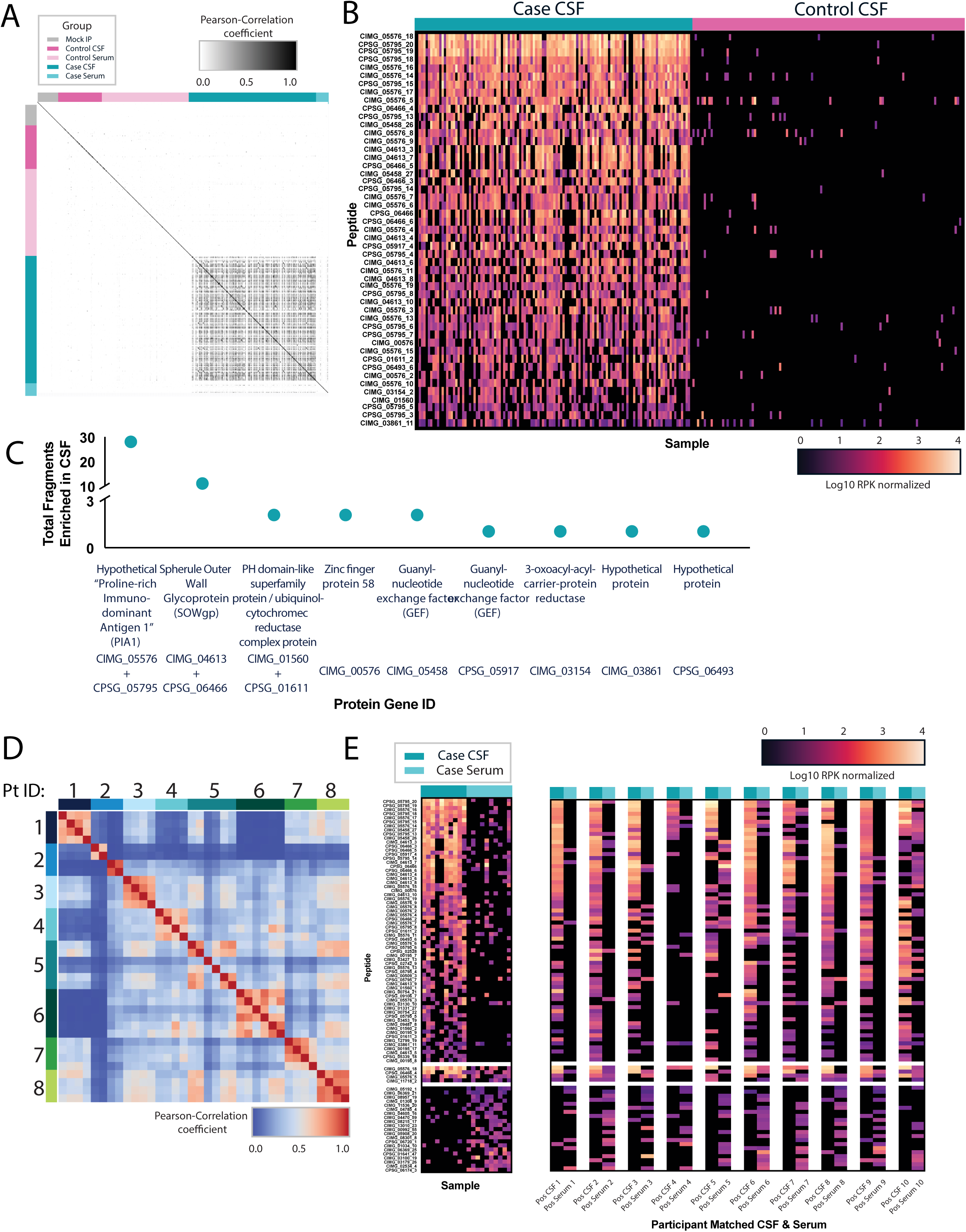
Compartment-specific antibody reactivity in CSF and serum from patients with coccidioidal meningitis. **(A)** Pairwise Pearson correlation matrix of log-transformed RPK values demonstrating strong within-cohort concordance among CM CSF samples. 43/163 representative OND samples shown as Control CSF. **(B)** Side-by-side heatmaps of RPK values showing peptide enrichment across CM CSF samples (left) and 108/163 representative OND control CSF samples (right); peptide enrichment is markedly higher in CM samples (p < 0.01). **(C)** Protein-level aggregation of enriched seroreactive peptides identifies 49 peptides derived from 9 *Coccidioides* proteins, with SOWgp and PIA1 representing the dominant immunogenic targets. **(D)** Comparison of matched CSF and serum samples (n=10 participants). Of 69 total CSF-enriched seroreactive peptides, 65 (94%) were detected exclusively in CSF and 4 (6%) were shared with serum; of 25 total serum-enriched seroreactive peptides, 21 (84%) were detected exclusively in serum. Shared peptides were primarily derived from SOWgp fragment 4 (AA150-200) and PIA1 fragments 5 and 18 (AA175-225 and AA500-550, respectively), while PIA1-derived peptides predominated among CSF-exclusive reactivities. **(E)** Longitudinal analysis of CSF samples from 7 participants. Within-participant pairwise Pearson correlation coefficients were consistently higher than between-participant coefficients for all 7 participants (mean within-participant r = 0.53-0.82).

Comparison of matched CM positive CSF and serum samples (n=10) revealed minimal overlap, with only ∼6% of enriched seroreactive peptides shared between compartments both within a given participant and across all 10 participants (**Figure 3D-E**). Shared reactivity was limited to a small subset of peptides derived from two immunodominant antigens: SOWgp and PIA1. Specifically, shared peptides included SOWgp fragment 4 (AA150-200), also seen in the Category 5 sera, and PIA1 fragments 5 and 18 (AA175-225 and AA500-550), respectively.

### Longitudinal CSF *Coccidioides* antibody dynamics in CM

To assess the similarity of PhIP-Seq reactivity profiles across longitudinal CSF samples, we computed pairwise Pearson correlation coefficients across samples from 7 participants with CM and visualized them as a clustered heatmap (**Figure 3D**). Within-patient correlations were consistently higher than between-patient correlations for all participants, with mean within-patient r values ranging from 0.53 to 0.82.

### Structural features of enriched antigenic peptides

We analyzed the structural characteristics of the consistently enriched seroreactive peptides identified by PhIP-Seq of CM-positive CSF. Antibody enrichment spanned much of the length of PIA1 and SOWgp (**Figure 4A**). Notably, these proteins contain proline-rich regions with repetitive sequence motifs (**Supplemental Table 5**).

**Figure 4:**
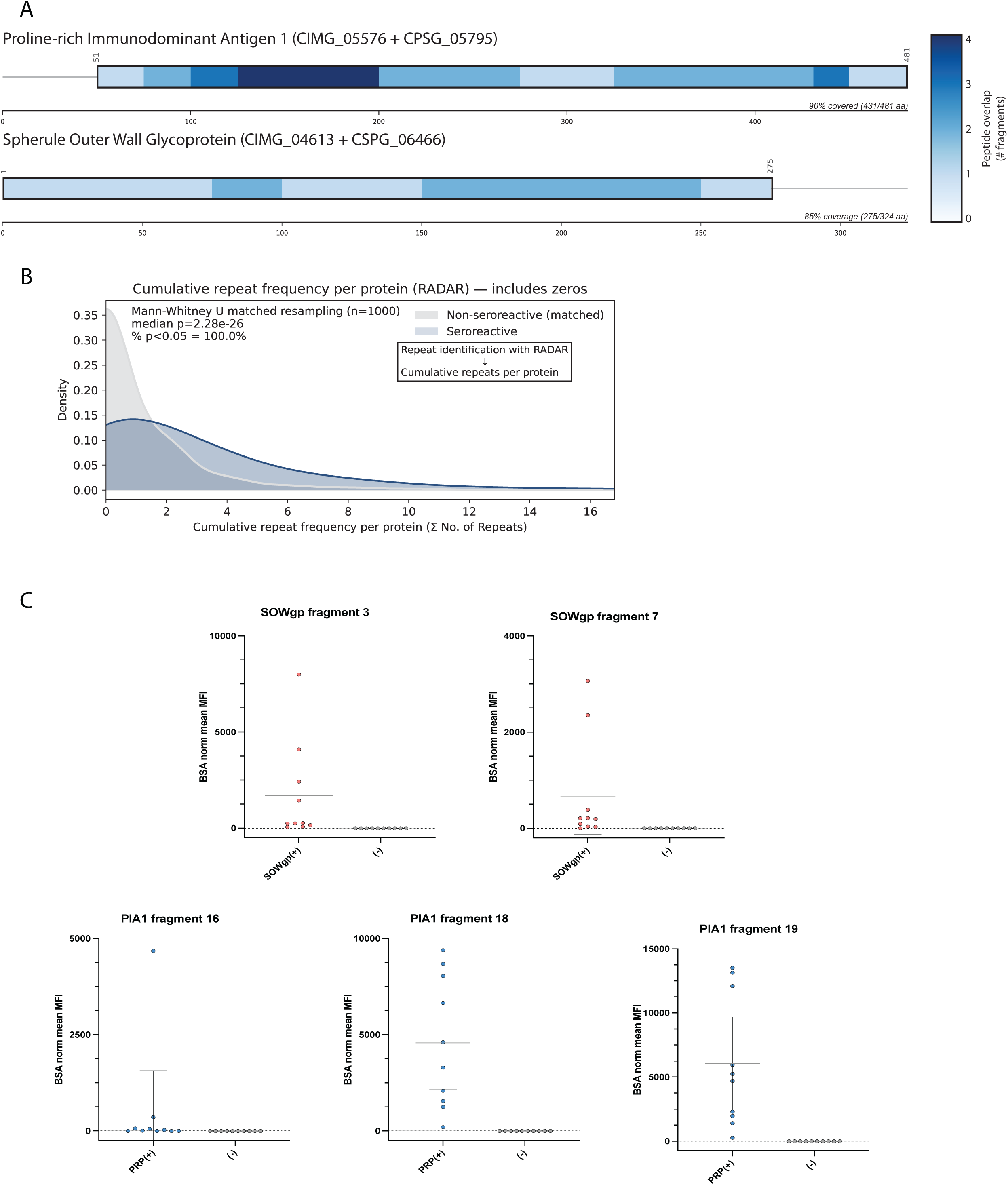
Structural characteristics and diagnostic validation of immunodominant *Coccidioides* antigens. **(A)** Distribution of antibody enrichment across the full length of PIA1 (CIMG_05576/CPSG_05795) and SOWgp (CIMG_04613/CPSG_06466). Peptides from both *C. immitis* and *C. posadasii* are mapped onto *the C. immitis* sequence, which serves as the reference for both proteins given the high similarity between orthologs. Enriched seroreactive peptides spanned 90% of the PIA1 and 85% of the SOWgp protein length (black box), with enrichment concentrated in proline-rich repetitive domains. **(B)** RADAR-based motif analysis comparing cumulative repetitive sequence element frequency between CSF-seroreactive proteins and non-seroreactive proteins. Seroreactive proteins exhibited significantly higher repeat content (Mann-Whitney U, median p = 2.28 × 10⁻² across 1,000 matched resampling iterations correcting for group size imbalance; seroreactive n=461, non-seroreactive n=10,622). **(C)** Luminex validation of five peptide antigens (SOWgp frag3, SOWgp frag7, PIA1-16, PIA1-18, PIA1-19) in CSF from coccidioidal meningitis (CM, n = 20) and other neurological disease controls (OND, n = 20), plotted as BSA-normalized MFI. For each antigen, CM samples (left) and OND controls (right) are shown, with the dashed line indicating the positivity threshold (MFI/background = 10).

Enriched seroreactive peptides (461 seroreactive vs. 10,622 non-seroreactive proteins) were significantly more likely to have a higher frequency of repetitive sequence elements relative to non-seroreactive peptides in the library (median cumulative repeat frequency: seroreactive vs. non-seroreactive, Mann-Whitney U, median p = 2.28 × 10⁻², 1,000 matched re-samplings). Motif analysis using RADAR identified recurring repeat structures across multiple enriched seroreactive peptides, consistent with prior studies^38^ suggesting that repetitive regions may be particularly immunogenic (**Figure 4B**).

### Orthogonal validation of PhIP-Seq detected *Coccidioides sp.* antigens using Luminex

Five antigens derived from SOWgp and PIA1 were selected for orthogonal validation in a Luminex-based assay:

PIA1_16 CINVGFPAGGLICHPACFPPSHQCPSGQQLRQRGGCSTCCEL, PIA1_18 CHDSDQPNPRPGDHDGPIIDHGALDPNHPHHHPHDSHQPN PIA1_19 CKPLKPKPKPESNPHDSDQPNPRPGDHDGPIIDHGALDPNH SOWgp_3 CDEYGYKMRKRGAKEHSYCDTYGCDGPMEPKPPKPTDCYG SOWgp_7 CKYGDCDYDDGYCDGPSKTSMKPEPPKPTDCYGDCKDGYD

All five peptides were synthesized at >95% purity, with an N-terminal cysteine to enable conjugation, and subsequently coupled to BSA at a 1:1 weight ratio (LifeTein LLC, Somerset, NJ). Of note, wild-type sequences were used, which contained internal cysteines that contributed to positional heterogeneity in the conjugation process.

We first validated these antigens on CSF from a subset of the CM CSF Discovery Cohort, consisting of participants with CM (n=20) and OND (n=20). Samples in this discovery subset were not reused in the subsequent validation cohort. Signal intensity varied across CM participants (**Figure 4C**), consistent with the heterogeneity of antibody responses observed in the PhIP-Seq datasets, and controls showed minimal signal. For SOWgp_3, specificity was 100% (95% CI: 83-100%), and sensitivity was 50% (95% CI: 27-73%). For SOWgp_7, specificity was 100% (95% CI: 83-100%), and sensitivity was 40% (95% CI: 19-64%). For PIA1_16, specificity was 100% (95% CI: 83-100%), and sensitivity was 55% (95% CI: 32-77%). For PIA1_18, specificity was 100% (95% CI: 83-100%), and sensitivity was 90% (95% CI: 68-99%). For PIA1_19, specificity was 100% (95% CI: 83-100%), and sensitivity was 90% (95% CI: 68-99%).

At the protein level, the two SOWgp antigens had a combined sensitivity of 55% (95% CI: 32-77%) and specificity of 100% (95% CI: 83-100%). The three PIA1 antigens had a combined sensitivity of 100% (95% CI: 83-100%) and specificity of 100% (95% CI: 83-100%). A combined antigen positivity score yielded perfect classification, with sensitivity of 100% and specificity of 100% (**Supplemental Table 6**).

Mean and standard deviation were calculated across all negative controls in the CM Discovery Cohort subset (n = 29) for each antigen-conjugated bead set. Negative control MFI/background ratios were consistently near 1 across all five bead sets (means 0.65-1.04, SDs 0.05-0.07), reflecting negligible non-specific binding above BSA background. Given this tight clustering near baseline, a fixed threshold of 10-fold signal over BSA background was chosen as the positivity cutoff. Positive CM control samples (n = 20) spanned a wide dynamic range (means 89-354, SDs 63-381 across bead sets), consistent with heterogeneous humoral responses across individuals and confirming that the signal threshold of 10-fold above BSA background captures true signal while remaining well above noise.

Next, Luminex Validation Cohort samples (34 CM samples from 32 participants and 36 OND samples from 36 participants) were randomized and plated by a second experimenter before processing by the first experimenter who was blinded to sample order and total number of positives. PIA1-derived peptides (PIA1_16, PIA1_18, and PIA1_19) combined achieved a sensitivity of 94% (95% CI: 80-99%) and a specificity of 100% (95% CI: 90-100%). Two samples without reactivity to PIA1 showed reactivity to SOWgp. SOWgp-derived peptides (SOWgp_3 and SOWgp_7) combined achieved a sensitivity of 59% (95% CI: 41-75%) and a specificity of 100% (95% CI: 90-100%). Combining the results of all five antigens, the sensitivity was 100% (95% CI: 90-100%) and the specificity was 100% (95% CI: 90-100%; Clopper-Pearson exact method) (**Supplemental Table 7**). Across the combined discovery and validation cohorts, which comprised non-overlapping participants (54 CM samples from 52 participants; 56 OND samples from 56 participants), the five-antigen panel achieved a sensitivity of 100% (95% CI: 93-100%) and a specificity of 100% (95% CI: 94-100%). These results confirm the diagnostic capability of a small set of *Coccidioides* antigens.

## DISCUSSION

Historically, identification of *Coccidioides* antigens relied on preparations of *coccidioidin* and *spherulin*, soluble antigen mixtures derived from culture filtrates of the saprobic (mycelial) and parasitic (spherule) phases, respectively. These complex antigen mixtures were characterized using two-dimensional immunoelectrophoresis (2D-IEP), which resolved dozens of antigenic components and established a reference framework for the fungal “antigenic mosaic.”^39^ Within this system, the most clinically relevant antigens were defined by their antibody reactivity: the tube precipitin (TP) antigen, associated with IgM responses in early infection, and the CF antigen, associated with later IgG responses. Subsequent biochemical and molecular studies identified these antigens as a β-glucosidase (TP antigen) and a chitinase (CF antigen), respectively.^9,10^

Here, we developed and applied a PhIP-Seq assay to assess the antigenicity of primarily linear antigens spanning all *C. immitis* and *C. posadasii* proteins in blood and CSF from *Coccidioides*-infected participants with a wide array of disease manifestations, ranging from asymptomatic exposure to DCM with CM. This approach identified both established antigens, such as SOWgp, and previously unknown antibody targets, including an uncharacterized protein CIMG_05576/CPSG_05795 that we refer to as PIA1. As the latter protein name suggests, we showed that the antigens we identified were disproportionately enriched with proline-rich and repetitive regions. Together, these findings expand the repertoire of *Coccidioides* antigens, offering a broader framework for understanding how antibody responses are organized across disease severity, anatomical compartments, and over time.

The enrichment of antibody binding to proline-rich and repetitive regions is notable. Similar structural features have been associated with immunodominant epitopes in other pathogens, in which sequence repetition may increase epitope density, accessibility, antibody binding kinetics, or B cell recognition.^38^ In the context of *Coccidioides*, these observations suggest that immunodominance may be shaped not only by antigen abundance or stage-specific expression, but also by underlying protein architecture. The proline-rich antigen PRA/Ag2 (CIMG_09696/CPSG_02952), one of the earliest characterized *Coccidioides* antigens, is similarly defined by its proline-rich composition,^40^ suggesting that this architectural feature recurs among immunodominant *Coccidioides* antigens.

While we identified antigens shared across participants ranging from asymptomatic *Coccidioides sp.* exposure to severe DCM, we also observed a narrowing of the antigenic repertoire with increasing disease severity. Participants with subclinical coccidioidomycosis (Category 1) mounted a more diverse humoral response in serum, with 55 enriched seroreactive peptides in serum, compared with 3-7 peptides in participants with more advanced disease. This narrowing may reflect an early, diverse antibody response that is lost or fails to develop in those who progress to DCM. Notably, the few peptides that persisted in participants with advanced disease mapped to likely cell surface or virulence-associated antigens: SOWgp (CIMG_04613/CPSG_06466), a surface adhesin implicated in host invasion, and PIA1 (CIMG_05576/CPSG_05795), an uncharacterized proline-rich protein identified as immunodominant in this study. The retention of reactivity to these two antigens even as the broader repertoire contracted suggests they represent the immunodominant core of the humoral anti-*Coccidioides* response, which is consistent with their diagnostic performance in participants with CM.

Many *Coccidioides spp.* antigens are spherule phase-specific since infection requires this phase for virulence. SOWgp is an immunodominant antigen expressed exclusively during the spherule stage. SOWgp is thought to function as an adhesin, facilitating the binding of spherules to host extracellular matrix proteins such as laminin and fibronectin to establish infection.^36^ To evade host defenses, the fungus secretes a metalloproteinase, Mep1, during its reproductive phase to digest SOWgp on the surface of endospores, effectively “stripping” them of this detectable antigen at their most vulnerable stage.^34^ As the sole spherule outer wall component recognized by antibodies in patients with DCM, SOWgp represents a promising but underutilized diagnostic target.^35,41^ In particular, its diagnostic potential in CSF has remained largely unexplored.

Prior antigen discovery work has demonstrated that immunization with select candidate antigens can reduce fungal burden and improve survival in murine models,^42–44^ suggesting that antibodies targeting surface-exposed or secreted proteins may contribute to protection. Subclinical participants who successfully clear infection may therefore harbor antibodies directed at spherule surface components critical to host invasion or immune evasion, responses that are narrowed or insufficient in individuals who progress to disseminated disease. While the Th1 response, including IFN-γ and IL-12, is well-established as critical for protection in murine coccidioidomycosis models,^45,46^ accumulating evidence suggests a complementary role for humoral immunity. B cells are necessary for full protection in vaccinated mice,^47^ and hypogammaglobulinemia has been associated with increased risk of DCM.^48^ Notably, however, Category 1 (unrecognized past infection) represents the smallest subgroup in our cohort, and the narrowed antigenic repertoire observed in participants with longer or more severe disease courses may reflect affinity maturation through somatic hypermutation rather than loss of protective breadth. Distinguishing these possibilities will require future studies with larger cohorts and longitudinal sampling across disease progression.

Our data demonstrate that the humoral immune response in CM is strongly compartmentalized. This compartmentalization may reflect sustained local antibody production by B cells recruited to or resident within the CNS, driven by *Coccidioides* that continues to proliferate in a site where antifungal penetration is reduced, while peripheral infection is more effectively controlled and the systemic response correspondingly wanes. This is consistent with the finding that the one patient in whom CSF and serum oligoclonal band analysis was performed was found to have evidence of intrathecal antibody production as evidenced by the presence of unique oligoclonal bands in the CSF. The most CNS-specific immunogenic protein was PIA1. We orthogonally validated this compartment-specific signal by developing a CSF-based Luminex assay using five CSF-specific SOWgp and PIA1 antigens. Although the peptides had individual sensitivities ranging from 40 to 90%, they were all 100% specific compared to a variety of CSF OND control samples with diseases that can mimic CM. As a multiplex serology, the combined sensitivity and specificity were 100 and 100%, respectively. In an expanded, blinded validation cohort spanning three independent sources (UCSF clinical CSF, UCD, and the UCSF NID biorepository; 34 CM and 36 OND CSF samples), the five-antigen panel reproduced 100% sensitivity and 100% specificity. Two CM samples were negative for all three PIA1 antigens yet were correctly classified by SOWgp reactivity, lowering standalone PIA1 sensitivity to 94% while the combined panel retained complete sensitivity, reinforcing that SOWgp and PIA1 capture non-overlapping components of the CSF antibody response.

The high degree of specificity of the CM-specific antigens for discriminating between CM participants and participants with other types of fungal meningitis is consistent with the fact that these antigens showed no meaningful sequence homology to proteins from other fungal pathogens. Together, these findings demonstrate that the CSF antibody response in CM is both compartment-specific and pathogen-specific, supporting its utility as a diagnostic assay.

The translational significance of these findings is twofold. First, it shows that proteome-wide antibody discovery is reduced to a practical, targeted platform that offers a high-value diagnostic yield.^28,49^ Second, it demonstrates newly identified antigens, particularly PIA1-derived targets, adding information beyond historically recognized antigens such as SOWgp. CF remains the clinical benchmark for CSF antibody detection, but it is technically demanding, slow, and imperfectly sensitive. In contrast, Luminex is a clinically validated platform that offers scalability, standardization, and the ability to measure multiple antigens simultaneously. In the validation cohort, eight CM samples had low CSF CF titers (1:2), near the lower limit of conventional CF reactivity, and all eight were correctly classified by the five-antigen panel. Notably, in the discovery cohort, our assay detected CM in samples with CF titers as low as 1:1, the lower limit of CF positivity. The lack of tight correlation between representative SOWgp and PIA1 peptide responses again supports that these targets are not merely redundant measurements of tightly coordinated antibody responses but instead capture distinct aspects of humoral immunity.

This study has several limitations. First, as with all peptide-based phage display approaches, the library primarily displays linear epitopes and may miss conformational and/or post-translationally modified antigens. This may contribute to why peptides from β-glucosidase (TP), chitinase (CF), and PRA/Ag2 were not among the enriched antigens. Second, several analyses remain limited by cohort size, especially within certain clinical subgroups, restricting the strength of inferences about disease severity-specific signatures. Third, sample collection occurred across variable timepoints relative to diagnosis and anti-fungal treatment, each of which may have influenced both the magnitude and composition of measured antibody responses. Fourth, the biological functions of several newly identified antigens, including PIA1, remain unknown. Although these targets appear diagnostically informative, their roles in pathogenesis, immune evasion, or host protection are unclear. PIA1 is a predicted secreted protein and is induced in the presence of macrophages, suggesting a role at the host-pathogen interface, but this remains to be tested directly.^50^ Lastly, although there was a limited number of patients in some of the subgroups (Category 1: unrecognized, past coccidioidomycosis infection; n=6), these samples represent rare clinical presentations for which specimens, particularly CSF, are difficult to obtain. Despite these limitations, the diversity of the cohort, the proteome-wide scope of the PhIP-Seq assay, and validation of the identified serological responses support the potential generalizability of these findings.

This study provides a broad antigenic map of humoral responses to *Coccidioides* infection and shows that such discovery can be translated into a targeted diagnostic platform. Importantly, the findings suggest that *Coccidioides* antibody responses encode information with anatomic and disease-stage specificity not captured by conventional assays. Future work should focus on further validating these targets in larger and more diverse cohorts, defining how treatment and timing influence these antibody signatures, and determining whether specific antigenic patterns are associated with fungal burden, prognosis, or protection. Beyond diagnostics, the expanded antigen repertoire identified here may also inform studies of host-pathogen interactions and vaccine design.

## Acknowledgements

We thank Jayant Rajan and Akshaya Ramesh for advice and guidance on computational phage library design. We thank the participants and their families for their participation in the study.

## Ethics statement

All human samples were obtained under institutional review board (IRB) approval at the University of California and collaborating institutions. All samples were de-identified prior to analysis.

## Data availability

PhIP-Seq data generated in this study will be made available for download at Dryad upon publication.

---

**Supplemental Table 1:**
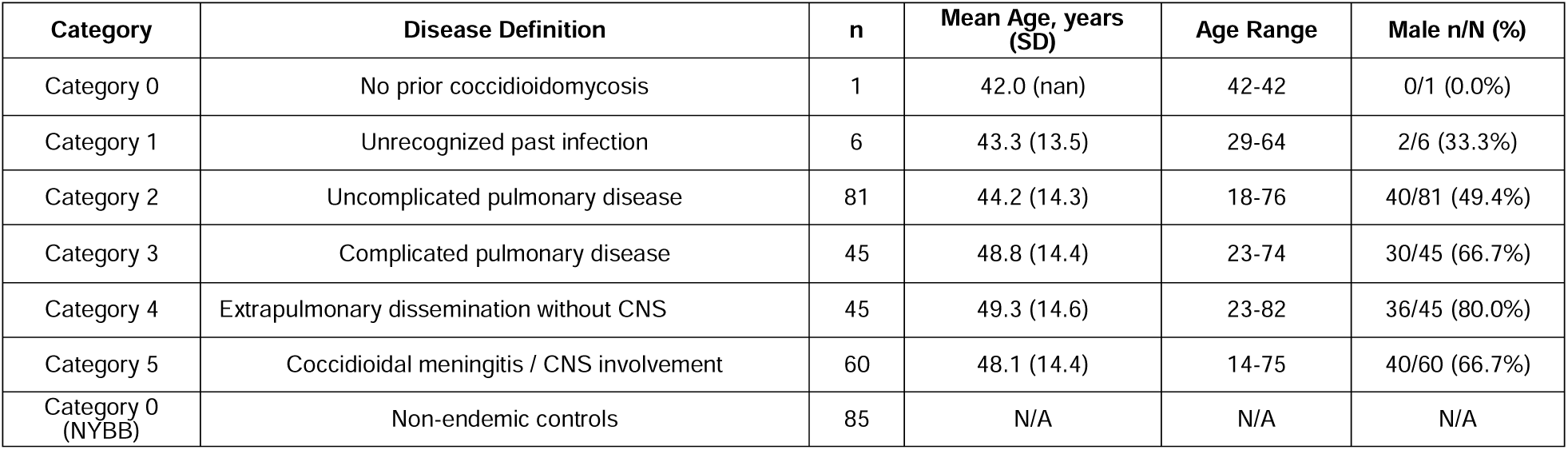
Demographic summary of the Coccidioidomycosis Subtype Serum Discovery Cohort (UCLA/Butte Lab, n=238; NYBB non-endemic controls, n=85). Disease categories assigned by infectious disease specialists.

**Supplemental Table 2:** Demographic summary of the CM CSF Discovery Cohort. . CM = coccidioidal meningitis; OND = other neurological disease. Geographic distribution shown for samples with available metadata.

| **Group** | **n** | **Mean Age, years (SD)** | **Age Range** | **Male n/N (%)** | **Geographic Distribution** |
| --- | --- | --- | --- | --- | --- |
| CM<br>(*Coccidioides*-positive CSF) | 108 | 45.4 (19.4) | 0-82 | 79/108 (73%) | CA (n=102); AZ (n=3); IN (n=1); WA (n=1) |
| OND<br>(*Coccidioides*-negative CSF) | 43 | N/A | N/A | 27/43 (63%) | CA (n=43) |
| OND (HIV associated cryptococcal meningitis) | 120 | 36 (N/A) | N/A | 70/120 (70%) | Uganda (n=120) |

**Supplemental Table 3:**
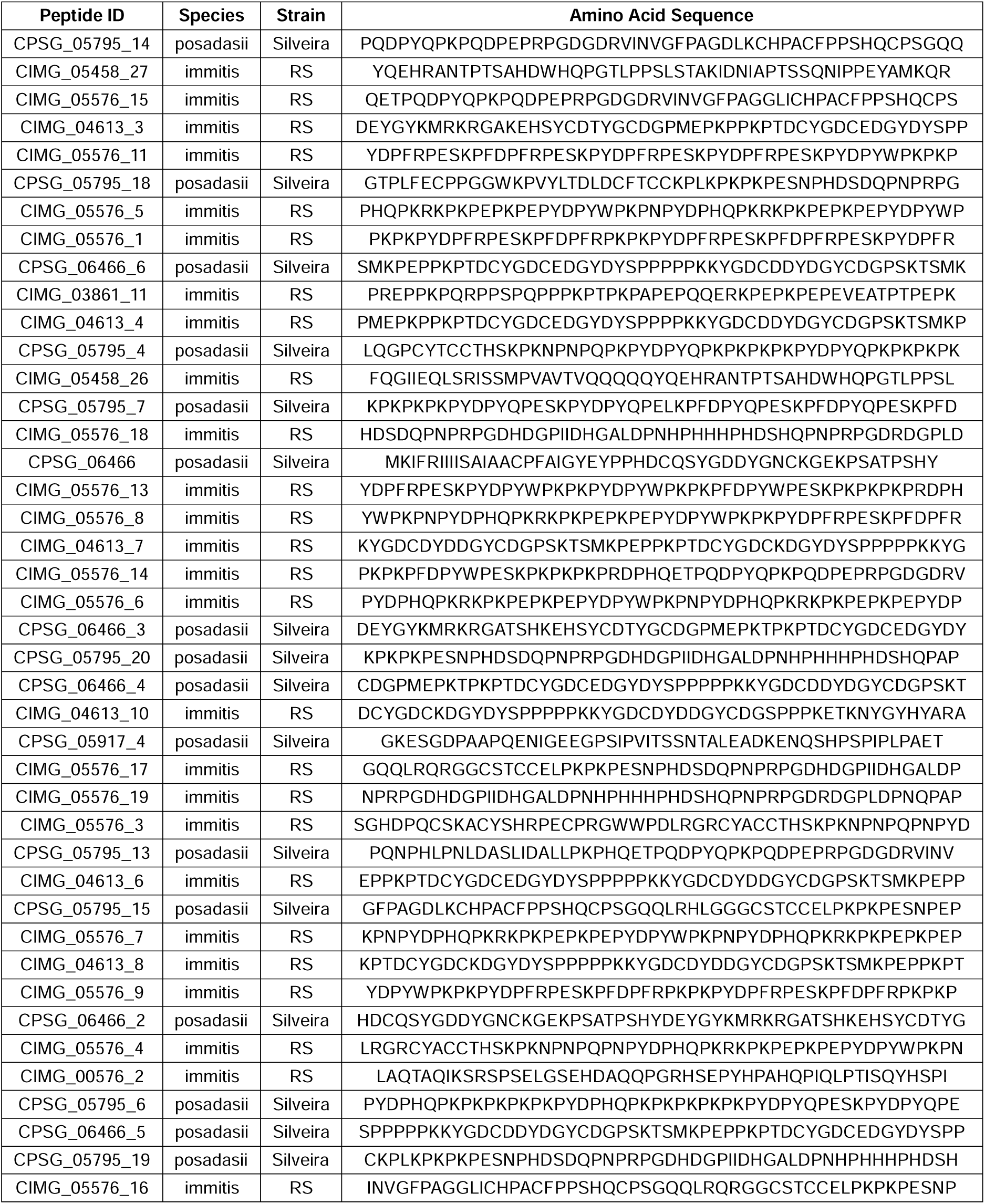
Enriched CSF peptides identified by PhIP-Seq analysis of the CM CSF Discovery Cohort. Peptides met enrichment criteria of ≥3-fold over mock-IP, Z-score ≥3, RPK ≥10, and presence in ≥50% of CM CSF samples. PIA1 = Proline-rich Immunodominant Antigen (CIMG_05576/CPSG_05795); SOWgp = spherule outer wall glycoprotein (CIMG_04613/CPSG_06466).

**Supplemental Table 4:** *Coccidioides* peptides enriched in CM CSF by PhIP-Seq, grouped by ortholog pairs (>95% amino acid identity). Enrichment criteria: ≥3-fold over mock-IP, Z-score ≥3, RPK ≥10, present in ≥50% of CM CSF samples.

| Ortholog Group<br>(>95% identity) | Genes | Total Peptides | protein |
| --- | --- | --- | --- |
| 1 | CIMG_05576 + CPSG_05795 | 28 peptides | Hypothetical protein ("Proline Rich Protein"PIA1) |
| 2 | CIMG_04613 + CPSG_06466 | 11 peptides | Spherule Outerwall glycoprotein |
| 3 | CIMG_05458 + CPSG_05917 | 3 peptides | Guanyl-nucleotide exchange factor |
| 4 | CIMG_00576 | 2 peptides | Zinc finger protein 58 |
| 5 | CIMG_01560 | 1 peptide | PH domain-like superfamily protein |
| 6 | CPSG_01611 | 1 peptide | ubiquinol-cytochrome c reductase complex protein |
| 7 | CIMG_03154 | 1 peptide | 3-oxoacyl-acyl-carrier-protein reductase |
| 8 | CIMG_03861 | 1 peptide | Hypothetical protein |
| 9 | CPSG_06493 | 1 peptide | Conserved hypothetical protein |

**Supplemental Table 5:**
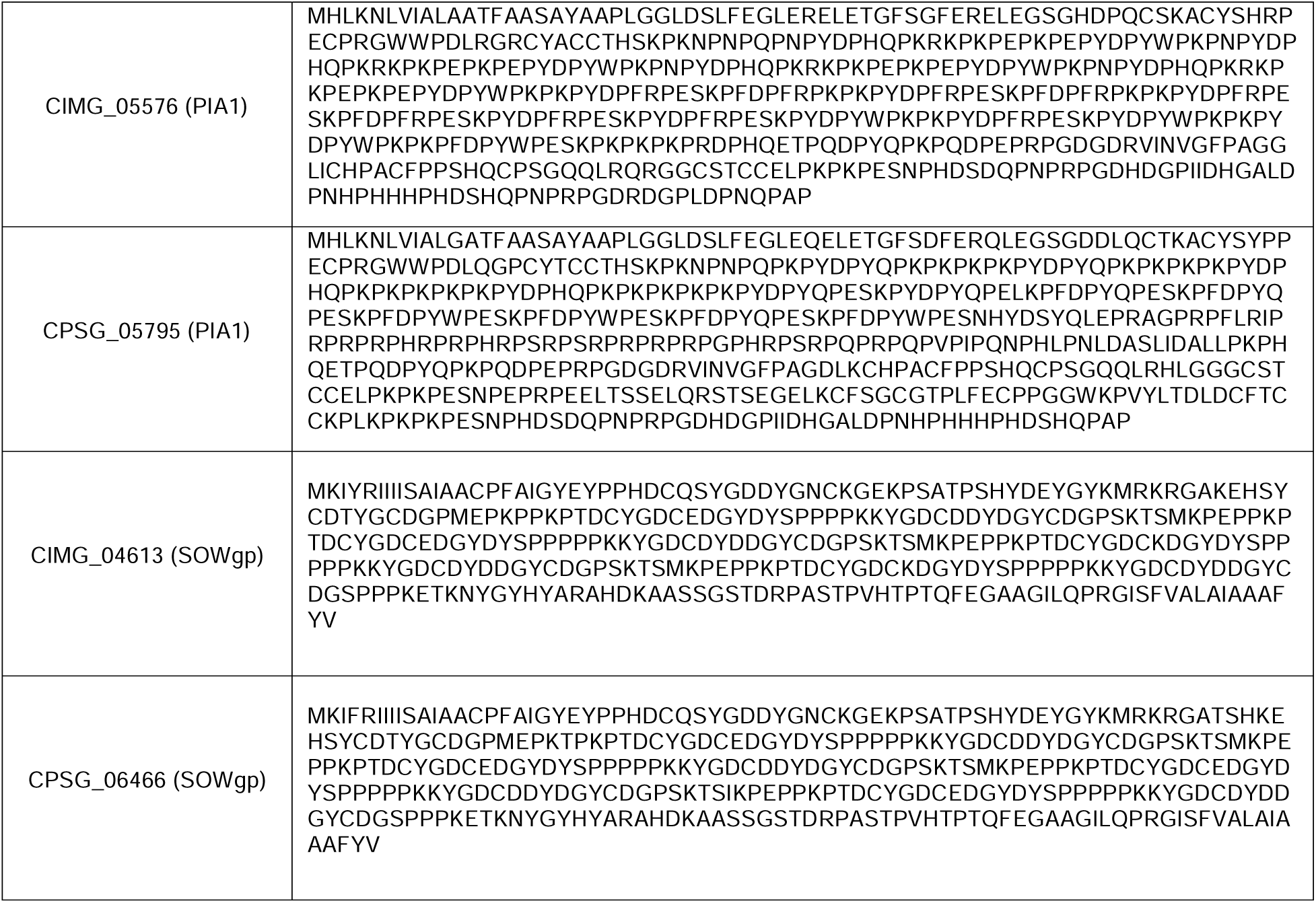
Proline-rich, repetitive sequences CIMG_05576, CPSG_05795, CIMG_04613, CPSG_06466.

**Supplemental Table 6:** Diagnostic Performance - CM Discovery Cohort. (n = 40),, CSF at 1:100 dilution, IgG secondary. Diagnostic performance of individual and aggregated antigens in the CM Discovery Cohort (CM n=20, OND n=20). PIA1-derived peptides demonstrated high sensitivity (PIA1_18 and PIA1_19: 90% [95% CI: 68-99%]) with 100% specificity (95% CI: 83-100%). Combined positivity across all three PIA1-derived antigens achieved 100% sensitivity and 100% specificity (95% CI: 83-100% for both; exact binomial [Clopper-Pearson] method). SOWgp-derived peptides showed lower sensitivity (40-55%) with 100% specificity.

| Test | Category | Sensitivity (%) | Specificity (%) | PPV (%) | NPV (%) | TP | FP | TN | FN |
| --- | --- | --- | --- | --- | --- | --- | --- | --- | --- |
| SOWgp fragment 3 | Individual Antigen | 50 | 100 | 100 | 66.7 | 10 | 0 | 20 | 10 |
| SOWgp fragment 7 | Individual Antigen | 40 | 100 | 100 | 62.5 | 8 | 0 | 20 | 12 |
| PRP fragment 16 | Individual Antigen | 55.0 | 100 | 100 | 69.0 | 11 | 0 | 20 | 9 |
| PRP fragment 18 | Individual Antigen | 90 | 100 | 100 | 90.9 | 18 | 0 | 20 | 2 |
| PRP fragment 19 | Individual Antigen | 90 | 100 | 100 | 90.9 | 18 | 0 | 20 | 2 |
| SOWgp (protein) | Protein Score | 55.0 | 100 | 100 | 69.0 | 11 | 0 | 20 | 9 |
| PRP (protein) | Protein Score | 100 | 100 | 100 | 100 | 20 | 0 | 20 | 0 |
| Combined Antigen | Combined Score | 100 | 100 | 100 | 100 | 20 | 0 | 20 | 0 |

**Supplemental Table 7:**
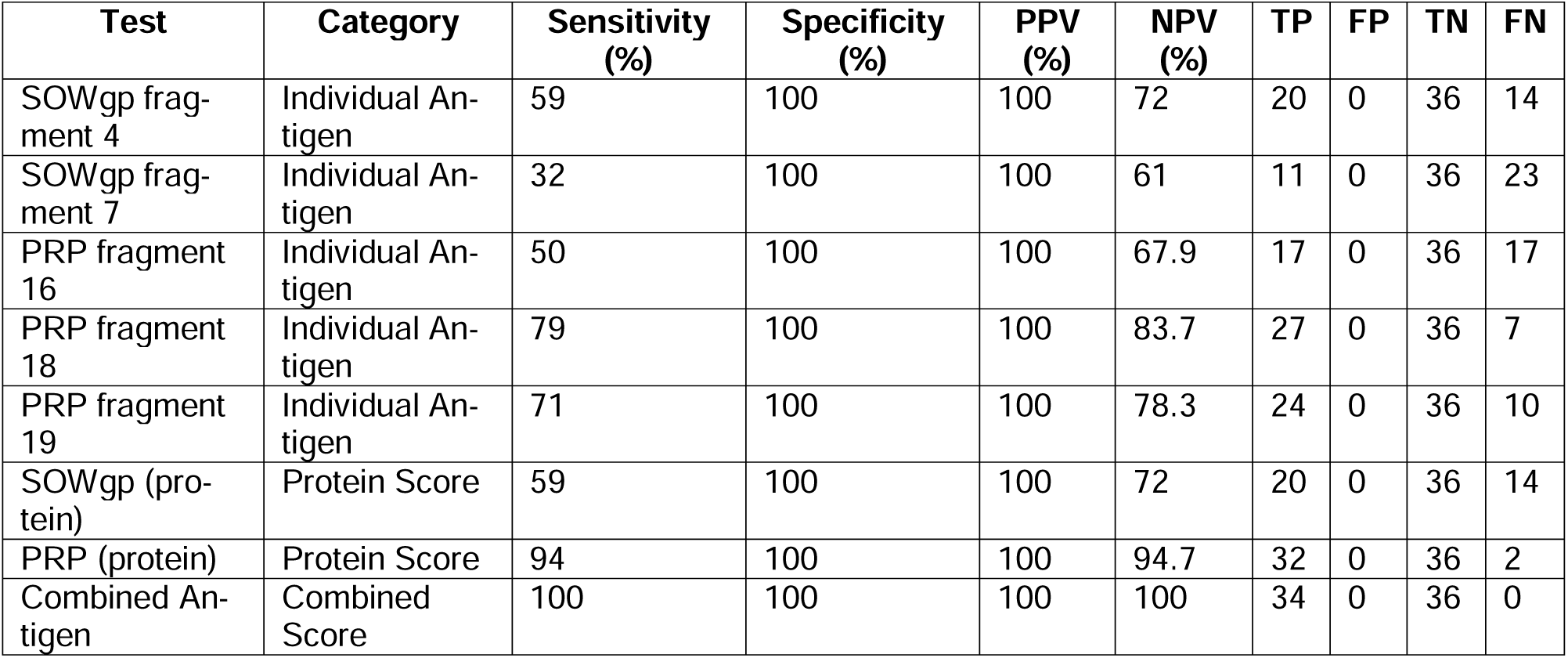
Diagnostic Performance — CM Validation Cohort (n = 70). Blinded experiment: CSF at 1:100 dilution, IgG secondary. Blinded CM validation cohort (CM n=32 participants / 34 samples; OND n=36 participants / 36 samples), confirming reproducibility of diagnostic performance. Combined five-antigen scoring achieved 100% sensitivity and 100% specificity (95% CI: 90-100% for both; Clopper-Pearson exact method). PIA1-derived peptides combined achieved 94% sensitivity (95% CI: 80-99%), while SOWgp-based detection remained less sensitive (59% [95% CI: 41-75%]).

